# Protocol for a Modified Delphi Consensus Study on the Clinical Assessment of Children with Suspected Olfactory Dysfunction

**DOI:** 10.64898/2026.07.01.26357062

**Authors:** Eishaan K. Bhargava, Katherine L. Whitcroft, Janine Gellrich, Gadi Fishman, Evan J. Propst, Laiquan Zou, Thomas Hummel

## Abstract

**Background:** Olfactory dysfunction (OD) affects approximately 22% of the general population, yet children account for only around 2% of specialist olfactory clinic attendees — a striking disparity that likely reflects systematic under-detection rather than lower disease prevalence. There are currently no internationally endorsed clinical guidelines governing the assessment of children with suspected OD. A recent international survey of 167 paediatric otolaryngologists across 36 countries found that more than half never perform psychophysical olfactory testing, a majority routinely order neuroimaging, and that multidisciplinary collaboration is largely lacking - findings that collectively motivate this consensus effort.

**Materials and Methods:** This is a prospectively registered, multi-round modified Delphi consensus study. A steering committee of seven international experts spanning paediatric otorhinolaryngology, chemosensory science, neuropaediatrics, and research methodology will generate an initial set of candidate consensus statements across 12 pre-defined clinical domains, derived from a structured narrative evidence review and practice-gap data. A panel of 15-25 international experts will vote anonymously over two to three sequential rounds using a 9-point Likert scale. Consensus is defined a priori as a median score ≥7 with ≥75% of panellists rating 7–9 and an IQR of ≤2. A post-Delphi virtual ratification meeting will follow voting rounds. Patient and Public Involvement (PPI) is embedded throughout: a family advisory group of 4-6 parents, caregivers, and young people with lived experience of paediatric OD, recruited in partnership with anosmie.org, will review candidate statements before Round 1, and PPI findings will inform statement framing before expert voting begins. This study was assessed using the Health Research Authority (HRA) online decision tool for England and found not to require NHS Research Ethics Committee (REC) review. The study will be conducted in accordance with the Research Governance Framework for Health and Social Care (2nd Edition).

**Discussion:** The final consensus statements and supporting methodology will be submitted for peer-reviewed publication. The study will be reported in compliance with the ACCORD and DELPHISTAR reporting standards for consensus methods. The protocol has been pre-registered prospectively on the Open Science Framework (OSF) prior to data collection.

## Introduction

### Burden of Olfactory Dysfunction in Children

Olfactory dysfunction (OD) in children is significantly under-recognised and understudied. While OD affects approximately 22% of the general population(1), only approximately 2% of all patients attending specialist olfactory clinics are under 18 years old(2). Children frequently fail to report olfactory impairment spontaneously, parents may not recognise its significance, and primary care clinicians often lack awareness of how to screen for or investigate suspected OD. The result is delayed diagnosis, missed syndromic associations, and limited access to support or intervention. The consequences for development, nutrition, safety awareness, and psychosocial well-being are substantial.(3) Despite a growing body of literature on paediatric OD, there are currently no specific international consensus recommendations governing how children with suspected OD should be assessed.

A scoping review by Payandeh et al. identified more than forty potential aetiologies of paediatric OD and highlighted the predominance of sensorineural mechanisms, while emphasizing the urgent need for validated age-appropriate olfactory tests and consistent clinical pathways.(4) Existing clinical review articles propose structured diagnostic approaches, but these represent expert opinion from individual groups rather than formally derived international consensus.(2,5) Stankevice et al. reinforced that diagnostic challenges are compounded by poor awareness and the significant impact of smell and taste loss on a child’s well-being.(6)

A recent international cross-sectional survey of paediatric otolaryngologists across 36 countries by Spencer et al has characterised global practice patterns in paediatric OD care.(7) The findings of this study directly motivate the present consensus effort and are summarised in Box 1.

#### Box 1.

Key global practice-pattern findings.

- 83% of paediatric ENT clinicians report seeing children with OD, yet 95% see fewer than ten such patients per year.
- 54.8% never perform psychophysical olfactory testing; routine testing (>75% of cases) is reported by only 15.3% of respondents.
- Testing frequency increases significantly with patient age (Cochran’s Q p<0.001), with minimal testing in children aged 0–2 years (0.6%) and 3–6 years (8.4%).
- 88.4% routinely order cross-sectional imaging when OD is the presenting symptom - a striking prioritisation of structural over functional assessment.
- Top barriers to objective testing: insufficient training (44.3%), time constraints (29.9%), institutional funding (28.1%).
- Multidisciplinary collaboration is negligible: dietitians involved routinely in only 1.7% of syndromic OD cases; speech-language pathology 5.8%; neuropsychology 2.6%; clinical genetics 21.4%; paediatric neurology 15.5%.
- Lack of referral guidelines was the most frequently cited barrier to collaboration (41.9%), without significant regional variation - indicating a widespread, systematic problem.
- Olfactory training offered in chronic OD by only 35.9% of respondents globally despite emerging evidence supporting efficacy.
- Respondent-identified advocacy priorities: better residency training (44.3%), integration of olfactory screening in primary care (32.9%), and national smell screening guidelines (31.7%).
- The authors conclude that standardised clinical guidelines, age-appropriate validated assessment tools, and formal interdisciplinary care pathways are urgently needed.

These findings establish three critical evidence-based justifications for the present consensus study: First, the absence of standardised guidelines is the principal structural driver of inconsistent practice, explicitly identified as the top priority for intervention by global respondents. Second, psychophysical testing is systematically under-utilised in favour of imaging, indicating a potential clinical-culture gap that consensus-derived recommendations on age-stratified test selection can directly address. Third, multidisciplinary collaboration has failed to emerge organically across health systems and will likely require explicit, expert-endorsed care-pathway recommendations to become routine. This creates a compelling case for a structured, internationally endorsed consensus document to guide clinical practice across both expert and non-expert centers.

### The Modified Delphi Approach

The Delphi method is increasingly used in medicine to develop consensus in clinical areas lacking robust evidence, providing structured expert guidance while determining shared principles adaptable to the individual patient. The International Paediatric Otolaryngology Group (IPOG) is an international organization that fosters collaboration among paediatric otolaryngology professionals worldwide and publishes consensus and guideline papers for both common and complex disorders.(8) IPOG has an established track record of using the modified Delphi method to generate consensus on topics where evidence is limited.(9–12) This protocol describes a methodologically rigorous modified Delphi consensus process for developing clinical recommendations on the assessment of children with suspected OD, reported in accordance with ACCORD(13) and DELPHISTAR(14) reporting standards.

### Aims and Objectives

The primary aim of this study is to develop an internationally endorsed consensus statement on the clinical assessment of children with suspected OD, using a modified Delphi process. The specific objectives are:

1. To define the minimum and comprehensive clinical history and examination elements relevant to paediatric OD assessment.
2. To establish consensus on age-appropriate psychophysical olfactory test selection.
3. To agree on indications for ancillary investigations – including neuroimaging, electrophysiology, and blood investigations.
4. To define multidisciplinary referral triggers, care coordination responsibilities, and complex case management pathways.
5. To establish consensus on follow-up standards, monitoring intervals, and safety and family education elements.

## Materials and Methods

### Overview and design

This is a prospectively registered, multi-round modified Delphi consensus study, consistent with the most recent IPOG precedents and with published guidelines for designing and conducting Delphi consensus studies in medical specialties.(15,16) The study will proceed through sequential phases (Figure 1):

- Phase 0: Infrastructure, steering committee orientation, protocol registration.
- Phase 1: Structured evidence review and initial statement generation.
- Phase 2: Expert panel identification and onboarding.
- Phase 3: Delphi voting rounds (2–3 rounds).
- Phase 4: Post-Delphi virtual consensus meeting and statement ratification.
- Phase 5: Manuscript preparation and submission.

**Figure 1:**
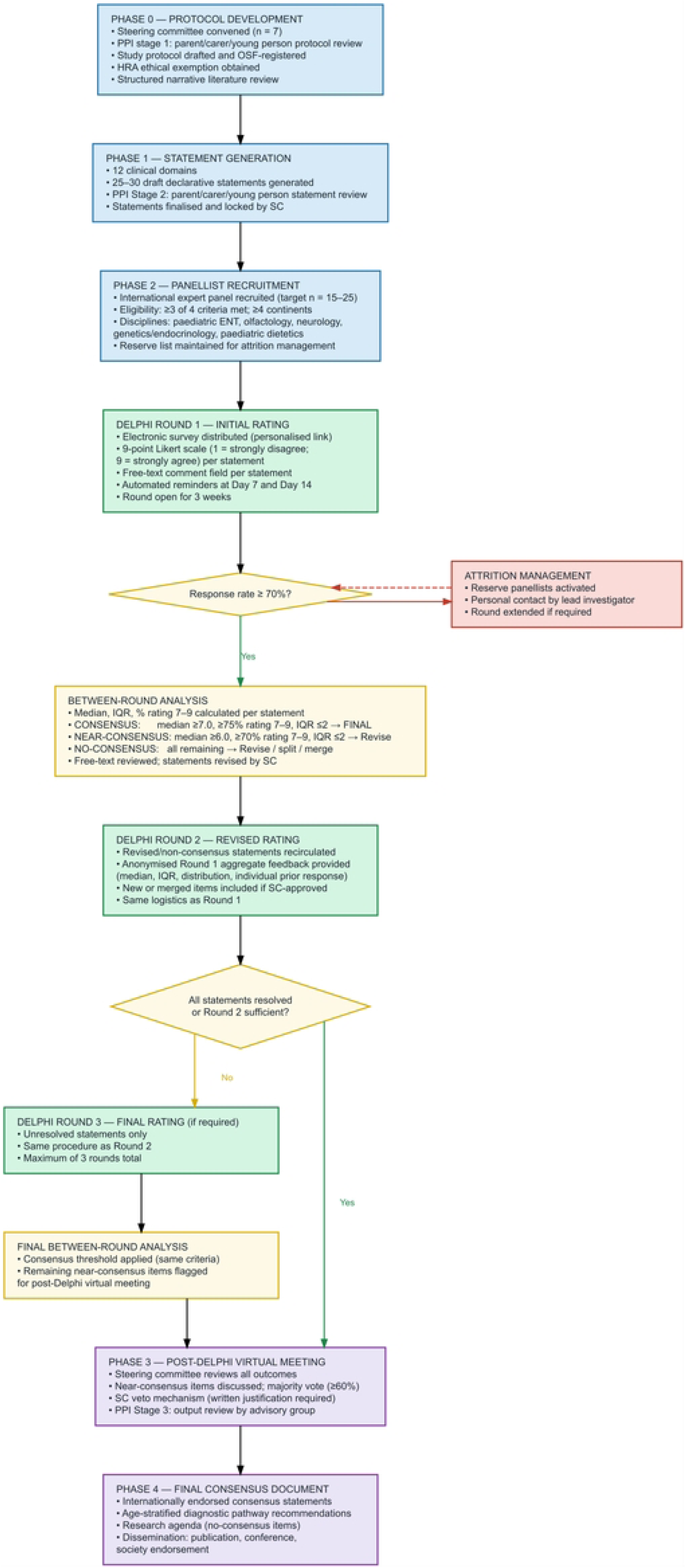
Overview of the modified Delphi consensus study.

### Phase 0: Pre-Study Infrastructure

#### Steering Committee

The steering committee comprises seven members with complementary expertise across paediatric otorhinolaryngology, chemosensory science, neuropaediatrics, and research methodology (Table 1).

**Table 1.** Steering committee composition.

| **Member** | **Institution** | **Country** | **Primary Expertise** |
| --- | --- | --- | --- |
| **Eishaan K. Bhargava (Lead)** | Sheffield Children's Hospital / University of Sheffield | UK | Paediatric ENT; paediatric chemosensory disorders |
| **Katherine L. Whitcroft** | Sheffield Children's Hospital / University of Sheffield | UK | ENT; Olfactory dysfunction |
| **Thomas Hummel** | TU Dresden | Germany | Chemosensory science; psychophysical testing |
| **Janine Gellrich** | TU Dresden | Germany | Paediatric neurology; paediatric OD |
| **Gadi Fishman** | Tzafron Medical Centre / Bar-Ilan University | Israel | Paediatric otolaryngology |
| **Evan J. Propst** | SickKids / University of Toronto | Canada | Paediatric otolaryngology |
| **Laiquan Zou** | Southern Medical University, Guangzhou | China | Psychology; chemosensory science |

#### Structured Evidence Review

A structured narrative review of existing literature on paediatric OD assessment will be conducted as the evidence base for initial statement generation (search strategy available for review as supplementary material S1).(17) The review will cover: Clinical history components and minimum dataset; validated psychophysical olfactory tests for children (including Sniffin’ Sticks(18,19), U-Sniff(20,21), qU-Sniff(22), NIH Toolbox(23), pBOT-6(24), Paediatric Smell Wheel(25), C-Sniff(26)); patient/parent reported outcome measures (PROMs); imaging indications and diagnostic/prognostic information; electrophysiological testing (olfactory event-related potentials, EEG-based techniques); multidisciplinary referral pathways; follow-up and safety counselling frameworks.

#### Protocol Registration and Ethics

This protocol was assessed using the Health Research Authority (HRA) online decision tool for England(27) (assessed 29 May 2026; Supplementary material S2). The tool confirmed that this study does not require NHS Research Ethics Committee review, as it does not involve NHS patients or service users as research participants, does not involve prospective collection of tissue or personal information from past or present NHS service users, and does not meet any of the criteria for mandatory REC review outlined in Question Sets 1-4 of the HRA decision algorithm. The study will be conducted in accordance with the Research Governance Framework for Health and Social Care (2nd Edition) and the Declaration of Helsinki principles as applicable to non-interventional research.

This protocol has been pre-registered prospectively on the Open Science Framework (OSF; osf.io) prior to data collection, in accordance with DELPHISTAR requirements.(14) All panellists will provide explicit informed consent prior to Round 1 access. ICMJE-format conflict of interest (COI) declarations will be collected from all steering committee members and panellists at enrolment.

### Phase 1: Statement Generation

Initial candidate consensus statements will be generated from three integrated sources:

- <u>Literature review output:</u> declarative statements derived from the structured evidence review, formatted for Likert-scale voting.
- <u>Practice-gap mapping from Spencer et al.’s survey</u>(7): each of the priority areas identified in the survey - awareness and education; screening and early identification; interdisciplinary collaboration; validated paediatric assessment tools; and guideline development - will be mapped directly to candidate consensus statements in this protocol’s domains.
- <u>Steering committee iterative drafting</u>: refinement through two virtual meetings, followed by PPI advisory group review prior to statement lock.

Statements will be grouped into 12 clinical domains (Table 2). A target of 25-30 carefully crafted statements is planned, with sufficiency defined by domain coverage rather than a fixed numerical target. Precedent IPOG consensus protocols have used 26–50 statements across comparable domain structures.(9–12)

**Table 2.** Clinical domains and indicative scope of consensus statements.

| Domain | Indicative scope |
| --- | --- |
| **1. Pre-referral and triaging** | Red flags triggering specialist referral; primary care screening; minimum referral information |
| **2. Clinical history** | Minimum dataset; onset (congenital vs. acquired); associated symptoms; family history |
| **3. Clinical examination** | Anterior rhinoscopy; nasal endoscopy; oral/craniofacial; neurological elements |
| **4. Psychophysical olfactory testing** | Tool selection by age band; minimum battery; cultural adaptation; norms; threshold vs. identification vs. discrimination components; retronasal testing indications/methods |
| **5. PROMs** | Patient-reported instruments; parental proxy reporting; age thresholds for self-reporting |
| **6. Taste and trigeminal function testing** | Indications; available methods; clinical scenarios |
| **7. Additional investigations** | Blood investigations for differential diagnosis; renal ultrasound indications |
| **8. Neuroimaging** | MRI indications (olfactory bulb volume, tract assessment, olfactory sulcus; shape of OB, shape of anterior skull base); sequencing |
| **9. Electrophysiology** | OERP indications; availability thresholds; clinical utility |
| **10. Multidisciplinary referral pathways** | Triggers (genetics, neurology, endocrinology, dietitian, SLT, psychology); MDT thresholds |
| **11. Follow-up and monitoring** | Minimum reassessment interval; components; escalation/discharge criteria |
| **12. Safety and family education** | Minimum safety counselling; caregiver education; aetiology-specific adaptations |

### Phase 2: Expert Panel Recruitment

#### Panel Size and Composition

A panel of 15–25 international experts will be recruited via email, with up to two reminders at fortnightly intervals. A minimum response rate of 70% per round will be required for validity. Steering committee members will not participate in voting rounds. Panellists will include: paediatric rhinologists and ENT surgeons with subspecialty interest in OD; olfactologists and chemosensory researchers with published paediatric experience; paediatric neurologists with experience in congenital anosmia or syndromic OD; clinical geneticists/endocrinologists with experience in Kallmann, CHARGE, or related conditions; and at least one paediatric dietitian to address the nutritional-care deficit identified by Spencer et al.(7)

#### Eligibility Criteria

Potential panellists must satisfy at least three of the following four criteria. This threshold permits inclusion of chemosensory researchers and basic scientists whose expertise is critical to statement development but who may not hold a direct clinical OD assessment role: (1) ≥2 years of clinical or research experience specifically involving paediatric OD or chemosensory disorders; (2) ≥1 peer-reviewed publication on paediatric OD, olfactory testing, or paediatric rhinology; (3) current membership of a relevant professional society (IPOG, CEORL, ARS, ISCS, ECRO, or national/international equivalent in their specialty); (4) active clinical practice at a centre that regularly assesses children with suspected OD.

#### Diversity Matrix

Panel composition will be internationally representative. Recruitment will actively target representation from at least four continents to ensure cultural validity of assessment recommendations, particularly regarding olfactory test cultural adaptation. A diversity matrix will capture: geographic region; practice setting; primary training background (paediatric ENT, adult rhinology with paediatric interest, chemosensory science, paediatric neurology, clinical genetics, dietetics); and experience.

### Phase 3: Delphi Voting Rounds

#### Survey Administration

Each round will be administered electronically. Each survey will be designed for completion within 30 minutes. Panellists will receive a personalised invitation link with a three-week voting window, with an automated reminder at two weeks. Surveys will be administered via Microsoft forms. Automated reminders will be issued at Day 7 and Day 14.

#### Voting Instrument

Each statement will be rated on a 9-point Likert scale (1 = strongly disagree; 9 = strongly agree), consistent with IPOG modified Delphi precedent.(9–12) A free-text comment field will accompany each statement. Panelist anonymity will be maintained throughout; individual responses will not be disclosed to other panellists. Following each round, descriptive statistics will be calculated for each statement: median score, interquartile range (IQR), and the proportion of panellists rating 7–9. These will be reported for all statements in the final manuscript’s supplementary materials. Between-round stability will be assessed by change in median and IQR; a shift of <0.5 in median and <1 point in IQR between successive rounds will be considered indicative of stable consensus. Free-text commentary will be synthesised thematically by the steering committee to inform statement revision. All analyses will be conducted using R Statistical Software (v4.5.3; R Core Team 2026).

#### A Priori Consensus Thresholds

Consensus categories are defined a priori (Table 3), aligning with broader literature suggesting 70–80% agreement as the standard for consensus.(16)

**Table 3.** A priori consensus threshold definitions.

| Category | Definition | Action |
| --- | --- | --- |
| Consensus | Median score $\geq 7.0$, $\geq 75\%$ panelists rating 7-9, IQR $\leq 2$ | Retained as a final consensus statement |
| **Near consensus** | Median score $\geq 6.0$, $\geq 70\%$ panelists rating 7-9, IQR $\leq 2$ | Retained for the subsequent round with revision informed by qualitative feedback |
| **No consensus** | All remaining statements | Substantially revised, split, merged, or removed if irresolvable after Round 3 |

#### Between-Round Statement Revision

Following each voting round, the steering committee will review results and apply the following decision rules: consensus statements are retained as final without further voting; near-consensus statements are revised based on qualitative feedback and redistributed; no-consensus statements are substantially revised, decomposed, merged, or removed; new statements proposed by panellists via free-text are reviewed by the steering committee for inclusion; a veto may be applied by majority steering committee decision when a consensus-threshold statement is found to be clinically unacceptable; vetoed statements become no-consensus items and written justification is provided to panellists in the subsequent round. Anonymised aggregate feedback (median score, interquartile range, score distribution, and each panellist’s own prior response) will be provided to panellists preceding each subsequent round.

#### Round Structure

In Round 1, the initial statement set will be distributed. Panellists will rate all statements and provide free-text commentary. In Round 2, revised and new statements (if any) will be redistributed with Round 1 anonymised aggregate scores. If required, Round 3 will be conducted for any revised or newly added statements following Round 2. There will be no further voting rounds beyond Round 3. Statements that have not achieved consensus after Round 3 will be carried forward to the Phase 4 virtual consensus meeting for resolution by structured discussion.

### Phase 4: Post-Delphi Consensus Meeting

A virtual consensus meeting of all panellists and steering committee members will be convened following voting rounds. The agenda will include: ratification of consensus statements; discussion and majority vote (defined as ≥60% agreement among attendees) on unresolved near-consensus statements – a lower threshold than the main consensus criterion is applied because synchronous discussion provides additional contextual information that may legitimately shift positions; agreement on wording; and framing of no-consensus items as research priorities.

### Patient and Public Involvement

A PPI advisory group of parents, caregivers, and young people with lived experience of paediatric OD has been assembled in partnership with anosmie.org, an international patient organisation for smell disorders with a significant French-speaking community.(28) PPI participants were asked to review the study summary and protocol documentation, and provided feedback via a bilingual (English/French) online questionnaire (Supplementary material S3) on study comprehension, domain prioritisation, lived-experience perspectives, and willingness for ongoing involvement. PPI feedback was collected during protocol development prior to OSF registration, consistent with best-practice guidance that PPI input should inform protocol design rather than follow it.

They will further review the draft candidate consensus statements before Round 1 voting. Their feedback – on completeness, clarity, relevance to lived experience, and potential harms from a patient perspective – will be reviewed by the steering committee before statements are finalised and distributed to the expert panel. PPI representatives will not participate in Delphi voting rounds, consistent with best-practice guidance separating expert clinical consensus from patient experience evidence.(29)

#### Stage 1 PPI Feedback Results

Six individuals completed the feedback questionnaire between 11 and 21 June 2026 via anosmie.org: four parents or carers of a child with olfactory dysfunction and two young people (aged ≥18) with personal lived experience. Children represented spanned three age groups (school age 7–11 years, n=3; adolescent 12–17 years, n=1; adult self-reporters, n=2). Five of six had a confirmed diagnosis; all reported congenital or suspected congenital anosmia. All respondents consented to future contact; all preferred French-language materials, with two also able to engage in English.

Study documents were rated highly for clarity across all three items (mean ratings out of 5: *what the research is about* 4.8; *why it matters* 4.7; *the respondent’s own role* 4.8; median 5 throughout). No section was identified as confusing.

The most frequently selected research priorities from a family perspective were Safety advice and education for families (4/6) and Testing taste and other sensations alongside smell (4/6), followed by Questionnaires about impact on daily life (3/6) – each corresponding directly to domains already represented in the protocol (Table 2, Domains 5, 6, 12).

Free-text responses identified five recurring concerns: (i) diagnostic delay and not being believed by clinicians; (ii) absence of post-diagnostic support or referral pathways; (iii) failure of primary and secondary care to recognise or refer – one family reached the correct pathway only via anosmie.org after three years of consultations; (iv) the profound impact of anosmia on eating, flavour, and social participation at mealtimes; and (v) the consistent neglect of psychological wellbeing, identity development, and interpersonal relationships in clinical assessments. Five of six respondents stated that children’s quality of life and emotional wellbeing are not currently adequately considered.

Mean interest in future PPI activities ranged from 3.5 to 3.8 out of 5. Preferred modalities were online questionnaire (n=5), email (n=4), and video call or online workshop (n=3 each).

Four of six respondents (67%) rated the research as “very useful - exactly the kind of research needed”; two (33%) as “useful - it will help, though some aspects could be strengthened”. Stage 1 feedback generated three recommendations for the steering committee prior to statement lock: (i) explicit coverage of psychological wellbeing and psychosocial development (including olfactory memory and identity) within the consensus domains; (ii) strengthened emphasis on nutritional impact and dietetic involvement (Domains 5 and 10); and (iii) inclusion of post-diagnostic support as an explicit component of Domain 12 (Safety and family education).

### Governance and Quality Assurance

#### Reporting Standards

The final manuscript will be reported in compliance with ACCORD(13) - a 35-item reporting checklist for consensus methods in biomedicine – and DELPHISTAR - the EQUATOR-registered Delphi reporting guideline.(14) A formal compliance checklist will be completed prior to submission; interim checklists are available for review as Supplementary material S4.

#### Conflict of Interest Management

ICMJE-format COI declarations will be collected from all steering committee members and panellists at enrolment. Declared conflicts will be summarised transparently in the methods and supplementary materials.

#### Attrition Management

Strategies to minimise panellist attrition include: survey design ≤30 minutes to completion; substantive aggregate feedback between rounds; automated reminders at Day 7 and Day 14; a minimum 70% response rate threshold per round; and a pre-identified reserve list of eligible panellists maintained for activation. If response rate falls below 70% in any round, reserve panellists will be activated, and attrition will be transparently reported and discussed in the final manuscript’s Limitations section.

#### Data Management

Data will be stored on a password-protected institutional server in accordance with UK GDPR data governance requirements. Panelist identities will be coded. Anonymised round data will be reported in supplementary materials.

## Discussion

### Rationale for a consensus approach

The absence of internationally agreed clinical guidance for the assessment of children with suspected OD represents a structural deficit with direct consequences for clinical practice. Modified Delphi consensus studies are among the most widely used and methodologically well-validated tools for generating practice guidance in clinical areas where randomised controlled trial evidence is absent or insufficient, and where practice variability is high. The modified Delphi approach - augmented by a post-Delphi ratification meeting – has been the standard framework employed by IPOG for consensus development in paediatric otolaryngology,(**9–12**) and is directly aligned with ACCORD(**13**) and DELPHISTAR(**14**) reporting standards. This methodological consistency with the IPOG precedent strengthens the legitimacy and potential adoption of the resulting consensus statement within the international paediatric ENT community.

### Strengths

Several methodological features strengthen the validity and potential impact of this study. The structured evidence base for statement generation, derived from a narrative review mapped against practice-gap data from Spencer et al.(**7**), ensures that consensus statements are anchored in the best available evidence and address the most clinically significant gaps in current practice. A priori definition of consensus thresholds and decision rules reduces the risk of post hoc manipulation of results, a key methodological safeguard. International representation across at least four continents ensures that the resulting recommendations will have cultural validity and global relevance, particularly regarding the selection of psychophysical olfactory tests with culturally adapted stimuli. Additionally, multi-disciplinary panel composition – spanning paediatric ENT, chemosensory science, paediatric neurology, clinical genetics, and dietetics – ensures that the 12 clinical domains are addressed with appropriate disciplinary breadth. Lastly, the integration of PPI through a structured family advisory group embeds patient and caregiver perspectives at the formative stage of statement development rather than as a post hoc consultation.

### Limitations

Modified Delphi consensus studies are subject to recognised limitations. Consensus reflects agreement among an expert panel at a specific point in time and is not equivalent to evidence derived from prospective studies or randomised trials. The selection of panellists inevitably introduces some degree of selection bias; mitigation through the diversity matrix and transparent eligibility criteria partially addresses, but does not eliminate, this limitation. Attrition between rounds is an inherent risk, particularly in international multi-round studies; the attrition management strategies described above are designed to minimise this. Steering committee veto, whilst essential as a clinical safety mechanism, introduces a degree of non-anonymous editorial control that departs from the pure Delphi model; this is handled transparently through the requirement for written justification of any veto. The absence of a paediatric patient voice in the Delphi voting rounds – a deliberate methodological decision to maintain separation between expert clinical consensus from patient experience evidence – means that the resulting statements may require implementation science research to evaluate their acceptability and feasibility from the family perspective. The PPI process described in this protocol partially addresses this gap, but full patient-partnered implementation research is beyond the scope of the present study. Future work should evaluate the acceptability and feasibility of implementing these consensus-derived recommendations from the perspectives of children and their families.

### Expected outputs and impact

The primary output of this study is an internationally endorsed consensus statement on the clinical assessment of children with suspected olfactory dysfunction, covering 12 clinical domains and ratified by a geographically and professionally diverse expert panel. This consensus statement is intended to provide the foundational evidence asset for a planned programme of larger, externally funded research, including the development and validation of age-stratified assessment tools, clinical trial design for olfactory training in paediatric populations, and health services research evaluating the feasibility of implementing consensus-derived pathways in routine clinical settings.

### Conclusions

This protocol describes a methodologically rigorous, internationally representative modified Delphi consensus study to develop the first expert-endorsed guidelines on the clinical assessment of children with suspected olfactory dysfunction. By combining a structured evidence base, a diverse international expert panel, transparent a priori consensus thresholds, and integrated patient and public involvement, the study is designed to produce recommendations with sufficient credibility, breadth, and clinical relevance to influence practice globally.

## Data Availability

This manuscript describes a study protocol no results data exist at the time of publication. All data generated during the conduct of the study — including anonymised aggregate Delphi voting data (per-statement median scores, IQRs, and proportion distributions per round) and the R analysis code — will be deposited in full on the Open Science Framework project page (https://osf.io/6gpsw/) upon submission of the results manuscript for publication. Individual panellist responses will not be shared publicly in order to preserve anonymity, consistent with the informed consent provided by participants aggregate round-level data will be made fully available without restriction.

## Acknowledgements

The authors gratefully acknowledge the support of anosmie.org and the members of the Patient and Public Involvement advisory group who reviewed the study documentation and provided feedback during protocol development. Their lived-experience perspectives have directly informed the scope and framing of this consensus study. We thank the families and young people who gave their time generously to contribute to this work.

## Methods statement

No data collection has yet commenced at the time of submission of this manuscript.

## Supplementary captions

**S1: Structured evidence review search protocol**

**S2: HRA online decision tool outcome**

**S3: Stage 1 PPI Feedback questionnaire**

**S4: ACCORD and DELPHISTAR checklists**

